# Predicting deseasonalised serum 25 hydroxy vitamin D concentrations in the D-Health Trial: an analysis using boosted regression trees

**DOI:** 10.1101/2020.08.23.20180422

**Authors:** Mary Waterhouse, Catherine Baxter, Briony Duarte Romero, Donald S. A. McLeod, Dallas R. English, Bruce K. Armstrong, Michael W. Clarke, Peter R. Ebeling, Gunter Hartel, Michael G. Kimlin, Rachel L. O’Connell, Hai Pham, Rachael M. Rodney Harris, Jolieke C. van der Pols, Alison J. Venn, Penelope M. Webb, David C. Whiteman, Rachel E. Neale

**Affiliations:** Population Health Department, QIMR Berghofer Medical Research Institute, Brisbane, Australia; Department of Endocrinology and Diabetes, Royal Brisbane and Women’s Hospital, Brisbane, Australia; Melbourne School of Population Health, University of Melbourne, Cancer Epidemiology and Intelligence Division, Cancer Council Victoria, Melbourne Australia; School of Public Health, University of Sydney, Sydney, Australia; Metabolomics Australia, Centre for Microscopy, Characterisation and Analysis, The University of Western Australia, Perth, Australia; Department of Medicine, School of Clinical Sciences, Monash University, Melbourne, Australia; Queensland University of Technology (QUT), School of Biomedical Sciences, Faculty of Health, Brisbane, Australia; NHMRC Clinical Trials Centre, University of Sydney, Sydney, Australia; School of Public Health, The University of Queensland, Brisbane, Australia; National Centre for Epidemiology and Population Health, College of Health & Medicine, The Australian National University, Canberra, Australia; Queensland University of Technology (QUT), School of Exercise and Nutrition Sciences, Faculty of Health, Brisbane, Australia; Menzies Institute for Medical Research, University of Tasmania, Hobart, Australia

**Keywords:** Vitamin D, randomized clinical trial, boosted regression trees, prediction model

## Abstract

**Background:** The D-Health Trial aims to determine whether monthly high-dose vitamin D supplementation can reduce the mortality rate and prevent cancer. We did not have adequate statistical power for subgroup analyses, so could not justify the high cost of collecting blood samples at baseline. To enable future exploratory analyses stratified by baseline vitamin D status, we developed a model to predict baseline serum 25 hydroxy vitamin D [25(OH)D] concentration.

**Methods:** We used data and serum 25(OH)D concentrations from participants who gave a blood sample during the trial for compliance monitoring and were randomised to placebo. Data were partitioned into training (80%) and validation (20%) datasets. Deseasonalised serum 25(OH)D concentrations were dichotomised using cut-points of 50 nmol/L, 60 nmol/L and 75 nmol/L. We fitted boosted regression tree models, based on 13 predictors, and evaluated model performance using the validation data.

**Results:** The training and validation datasets had 1788 (10.5% <50 nmol/L, 23.1% <60 nmol, 48.8 <75 nmol/L) and 447 (11.9% <50 nmol/L, 25.7% <60 nmol/L, and 49.2% <75 nmol/L) samples, respectively. Ambient UV radiation and total intake of vitamin D were the strongest predictors of ‘low’ serum 25(OH)D concentration. The area under the receiver operating characteristic curves were 0.71, 0.70, and 0.66 for cut-points of <50 nmol/L, <60 nmol/L and <75 nmol/L respectively.

**Conclusions:** We exploited compliance monitoring data to develop models to predict serum 25(OH)D concentration for D-Health participants at baseline. This approach may prove useful in other trial settings where there is an obstacle to exhaustive data collection.

## INTRODUCTION

The importance of vitamin D may extend beyond regulating calcium and phosphorus homeostasis. Numerous observational studies have found an inverse association between serum 25 hydroxy vitamin D [25(OH)D] concentration, a marker of vitamin D status, and risk of many diseases.^1^ Nonetheless, these results may be due to reverse causality or uncontrolled confounding; without high quality randomised trial evidence, we cannot confidently conclude that increasing serum 25(OH)D concentrations in the population through routine vitamin D supplementation or food fortification would improve health.

While randomised controlled trials (RCTs) of non-musculoskeletal outcomes have been performed,^2^ uncertainty of the benefit from supplementing vitamin D persists. A systematic review of meta-analyses and mostly small RCTs found that vitamin D supplementation may prevent premature all-cause mortality and mortality from cancer, and possibly reduces the risk of upper respiratory tract infections and asthma exacerbations.^2^ However, there was little evidence of a protective effect for most conditions.^2^ In addition, one large RCT has recently reported no benefit from vitamin D supplementation for incident cancer, cardiovascular disease, or all-cause mortality.^3^ These discrepancies with observational studies may be a consequence of inadequacies in trial designs, such as being under-powered, using a dose that is too low, or being of too short duration,^2^ or may reflect a true lack of benefit from vitamin D supplementation.

We launched the D-Health Trial^4^ in 2014 to determine whether supplementing older people (aged ≥ 60 years) with monthly doses of 60,000 international units (IU) of vitamin D over five years can prevent cancer and reduce the mortality rate. Our intention was to replicate what would happen if the Australian population was routinely supplemented with vitamin D, with no prior vitamin D screening. Consequently, we aimed to recruit a representative sample from the older Australian population.

Since our goal was not to determine the benefits of treating vitamin D deficiency, and we do not have statistical power for subgroup analyses by baseline vitamin D status, we could not justify the substantial cost (estimated at several millions of dollars) of measuring 25(OH)D in all participants at baseline. However, there is value in conducting exploratory analyses stratified by baseline vitamin D status, as any effect of supplementation may be more pronounced amongst people with lower serum 25(OH)D concentrations prior to supplementation.^5^

During the intervention phase of the D-Health Trial, we collected blood samples from a random sample of participants (approximately 20%) for the purpose of compliance monitoring. Here we describe how we used measured serum 25(OH)D concentration from the placebo group (while maintaining blinding), and participants’ self-reported data, to develop and validate a model to predict deseasonalised baseline serum 25(OH)D concentrations for the entire cohort. Our approach may be useful to others conducting trials for which prohibitive cost or other obstacles prevent the collection of data on a possible effect modifier.

## METHODS

### Trial overview

The D-Health trial is a double-blind, randomised, placebo-controlled trial of monthly high-dose vitamin D supplementation in older Australians. We have published detailed trial methods and baseline characteristics^4^ and a statistical analysis plan for the primary and secondary outcomes of the trial.^6^ Using the Australian electoral roll as a sampling frame, we invited 421,207 people aged 60-79 years to participate. We also sought volunteers. People were not eligible if they self-reported a history of osteomalacia, sarcoidosis, hyperparathyroidism, hypercalcemia, or kidney stones, or if they reported taking supplementary vitamin D at a daily dose exceeding 500 IU. We randomised 21,315 people in a 1:1 ratio to receive either 60,000 IU of cholecalciferol (vitamin D^3^) or placebo on the first day of each month for five years. Randomisation occurred between February 2014 and May 2015, and was undertaken within strata of age (60-64; 65-69; 70-74; 75+ years), sex, and state of residence (New South Wales, Queensland, South Australia, Tasmania, Victoria, Western Australia) at baseline. The primary outcome is all-cause mortality and the secondary outcomes are total cancer incidence and colorectal cancer incidence; these will be ascertained via future linkage to death and cancer registries. The QIMR Berghofer Medical Research Institute Human Research Ethics Committee approved the trial and all participants gave written or online consent to participate. All participants completed the intervention phase by 1^st^ February 2020.

### Data collection

#### Baseline survey

Participants completed a survey at baseline that included questions about factors that we had previously shown to predict serum 25(OH)D concentration.^7^ We used self-reported height and weight to calculate body mass index (BMI, kg/m^2^). To capture information on skin phenotype, we asked people to report how much their skin would tan if they spent several weeks outdoors in the sun in summer. We used data about living arrangements to derive an indicator of whether or not the person was living alone. Participants were asked to rate their quality of life (QoL) on a five-point scale, ranging from poor to excellent. We asked about smoking history, alcohol consumption and physical activity, which was used to calculate typical energy expenditure as metabolic equivalent tasks (METs) per week. Data on the amount of time spent outdoors between 9 am and 4 pm on each day of the previous week was used to estimate total hours of sun exposure. Participants were asked to document their use of vitamin D supplements and intake of vitamin D-rich foods (using a pre-specified list of vitamin D-rich foods) in the previous month. We used AUSNUT 2007 and food/supplement composition details supplied by manufacturers to calculate total daily intake of vitamin D from diet and supplements in IU. The groups into which we categorised these variables are shown in Table 1.

**Table 1.** Summary statistics for deseasonalised serum 25(OH)D concentration for all participants and by levels of predictor variables in the training and validation datasets.

| Predictor variable | Deseasonalised serum 25(OH)D concentration (nmol/L) |  |  |  |  |  |  |  |  |  |
| --- | --- | --- | --- | --- | --- | --- | --- | --- | --- | --- |
|  | N (%)<sup>1</sup> |  | Training dataset (N = 1788) |  |  |  | Validation dataset (N = 447) |  |  |  |
|  | Training | Validation | Mean (SD) | % |  |  | Mean (SD) | % |  |  |
|  |  |  |  | < 50 nmol/L | < 60 nmol/L | < 75 nmol/L |  | < 50 nmol/L | < 60 nmol/L | < 75 nmol/L |
| **Age (years)** |  |  |  |  |  |  |  |  |  |  |
| < 70 | 615 (34.4) | 166 (37.1) | 78.5 (23.3) | 10.1 | 21.3 | 46.0 | 80.3 (25.0) | 12.0 | 20.5 | 44.0 |
| 70 to < 75 | 494 (27.6) | 125 (28.0) | 77.9 (24.4) | 9.9 | 23.1 | 48.6 | 78.5 (22.8) | 13.6 | 24.0 | 46.4 |
| ≥ 75 | 679 (38.0) | 156 (34.9) | 75.9 (23.1) | 11.2 | 24.7 | 51.4 | 74.0 (22.9) | 10.3 | 32.7 | 57.1 |
| **Sex** |  |  |  |  |  |  |  |  |  |  |
| Men | 901 (50.4) | 223 (49.9) | 76.4 (22.9) | 10.2 | 23.9 | 51.8 | 76.5 (23.5) | 12.1 | 27.8 | 52.9 |
| Women | 887 (49.6) | 224 (50.1) | 78.3 (24.1) | 10.7 | 22.3 | 45.7 | 78.7 (24.0) | 11.6 | 23.7 | 45.5 |
| **Month** |  |  |  |  |  |  |  |  |  |  |
| January | 170 (9.5) | 29 (6.5) | 73.7 (20.8) | 11.8 | 22.9 | 55.9 | 74.7 (24.4) | 13.8 | 37.9 | 55.2 |
| February | 178 (10.0) | 36 (8.1) | 80.5 (24.5) | 7.3 | 21.3 | 42.7 | 76.0 (16.3) | 2.8 | 16.7 | 50.0 |
| March | 152 (8.5) | 47 (10.5) | 77.6 (24.9) | 11.2 | 23.0 | 47.4 | 76.1 (20.6) | 12.8 | 21.3 | 53.2 |
| April | 152 (8.5) | 38 (8.5) | 77.7 (27.9) | 12.5 | 25.7 | 52.0 | 69.7 (21.5) | 18.4 | 28.9 | 76.3 |
| May | 185 (10.3) | 50 (11.2) | 80.9 (25.3) | 9.2 | 23.2 | 42.2 | 82.6 (27.3) | 12.0 | 22.0 | 44.0 |
| June | 176 (9.8) | 31 (6.9) | 75.7 (26.2) | 15.9 | 27.3 | 54.0 | 76.6 (28.6) | 25.8 | 35.5 | 48.4 |
| July | 155 (8.7) | 49 (11.0) | 72.9 (23.3) | 15.5 | 29.0 | 54.2 | 75.0 (23.8) | 16.3 | 28.6 | 55.1 |
| August | 134 (7.5) | 30 (6.7) | 77.9 (21.3) | 9.0 | 23.9 | 45.5 | 74.9 (24.1) | 13.3 | 33.3 | 46.7 |
| September | 141 (7.9) | 45 (10.1) | 79.2 (21.5) | 8.5 | 16.3 | 47.5 | 81.6 (27.1) | 11.1 | 26.7 | 44.4 |
| October | 139 (7.8) | 40 (8.9) | 77.6 (22.2) | 9.4 | 23.0 | 48.2 | 83.7 (24.4) | 2.5 | 20.0 | 32.5 |
| November | 133 (7.4) | 33 (7.4) | 75.5 (19.6) | 7.5 | 21.1 | 49.6 | 80.2 (22.5) | 6.1 | 21.2 | 39.4 |
| December | 73 (4.1) | 19 (4.3) | 79.4 (18.9) | 2.7 | 15.1 | 43.8 | 77.6 (17.6) | 5.3 | 21.1 | 42.1 |
| **Ambient ultraviolet irradiation (J/m<sup>2</sup>)<sup>2</sup>** |  |  |  |  |  |  |  |  |  |  |
| < 1250 | 315 (17.6) | 84 (18.8) | 72.3 (24.5) | 17.1 | 31.7 | 58.1 | 73.6 (25.2) | 21.4 | 34.5 | 54.8 |
| 1250 to < 2500 | 437 (24.4) | 107 (23.9) | 79.7 (23.8) | 9.2 | 21.1 | 43.9 | 78.7 (27.1) | 14.0 | 27.1 | 49.5 |
| 2500 to < 3750 | 285 (15.9) | 81 (18.1) | 77.5 (24.7) | 11.2 | 23.9 | 50.5 | 78.9 (23.4) | 8.6 | 21.0 | 50.6 |
| 3750 to < 5000 | 262 (14.7) | 77 (17.2) | 77.6 (22.4) | 9.5 | 21.8 | 46.2 | 77.7 (20.6) | 6.5 | 24.7 | 46.8 |
| ≥ 5000 | 489 (27.3) | 98 (21.9) | 78.3 (22.2) | 7.4 | 19.6 | 47.4 | 78.6 (21.3) | 8.2 | 21.4 | 44.9 |
|  | N (%)<sup>1</sup> |  | Training dataset (N = 1788) |  |  |  | Validation dataset (N = 447) |  |  |  |
|  |  |  | Mean (SD) | % |  |  | Mean (SD) | % |  |  |
|  | Training | Validation |  | < 50 nmol/L | < 60 nmol/L | < 75 nmol/L |  | < 50 nmol/L | < 60 nmol/L | < 75 nmol/L |
| Physical activity (METs/week) |  |  |  |  |  |  |  |  |  |  |
| Low (< 18) | 350 (28.7) | 95 (30.4) | 77.1 (25.1) | 15.1 | 26.0 | 48.0 | 76.8 (25.5) | 15.8 | 27.4 | 45.3 |
| Moderate (18 to < 45) | 449 (36.8) | 107 (34.3) | 77.8 (21.4) | 7.1 | 20.9 | 46.1 | 80.9 (22.9) | 7.5 | 18.7 | 43.9 |
| High (≥ 45) | 420 (34.5) | 110 (35.3) | 80.6 (23.0) | 6.7 | 17.4 | 44.3 | 80.9 (26.0) | 10.9 | 21.8 | 45.5 |
| Time outdoors (hours/week) |  |  |  |  |  |  |  |  |  |  |
| < 5 | 292 (23.7) | 75 (24.1) | 76.7 (25.2) | 15.8 | 25.3 | 47.6 | 76.2 (25.2) | 14.7 | 29.3 | 50.7 |
| 5 to < 10 | 274 (22.3) | 52 (16.7) | 77.0 (21.6) | 9.1 | 22.3 | 46.7 | 79.0 (23.0) | 9.6 | 23.1 | 42.3 |
| 10 to < 15 | 237 (19.3) | 63 (20.3) | 76.9 (23.2) | 9.3 | 24.1 | 49.4 | 78.0 (23.2) | 9.5 | 20.6 | 47.6 |
| 15 to < 25 | 264 (21.5) | 59 (19.0) | 80.4 (23.1) | 6.8 | 17.8 | 44.3 | 82.9 (28.2) | 13.6 | 20.3 | 42.4 |
| ≥ 25 | 163 (13.3) | 62 (19.9) | 84.1 (22.9) | 3.1 | 13.5 | 39.9 | 82.6 (23.6) | 8.1 | 17.7 | 38.7 |
| Tan moderately or deeply |  |  |  |  |  |  |  |  |  |  |
| No | 593 (33.3) | 147 (33.3) | 74.7 (24.4) | 14.2 | 27.3 | 54.0 | 74.7 (22.7) | 15.0 | 29.3 | 52.4 |
| Yes | 1188 (66.7) | 295 (66.7) | 78.7 (23.1) | 8.7 | 21.0 | 46.4 | 79.1 (24.2) | 10.2 | 24.1 | 47.8 |
| Body mass index (kg/m²) |  |  |  |  |  |  |  |  |  |  |
| < 25 | 563 (32.6) | 129 (30.3) | 80.5 (24.9) | 9.6 | 20.2 | 42.8 | 81.8 (23.6) | 9.3 | 21.7 | 39.5 |
| 25 to < 30 | 717 (41.5) | 194 (45.5) | 77.3 (22.2) | 8.5 | 22.6 | 49.8 | 76.2 (23.5) | 11.3 | 26.3 | 52.6 |
| ≥ 30 | 449 (26.0) | 103 (24.2) | 73.2 (23.5) | 14.5 | 28.3 | 55.2 | 75.3 (23.2) | 13.6 | 27.2 | 55.3 |
| Alcohol consumption (drinks/week) |  |  |  |  |  |  |  |  |  |  |
| < 1.0 | 442 (25.8) | 136 (30.8) | 76.2 (24.8) | 11.3 | 25.3 | 53.4 | 74.6 (22.8) | 12.5 | 31.6 | 54.4 |
| 1.0-2.0 | 261 (15.2) | 72 (16.3) | 76.4 (21.5) | 9.2 | 23.4 | 49.4 | 75.8 (23.6) | 16.7 | 26.4 | 55.6 |
| 2.1-7.0 | 499 (29.1) | 114 (25.9) | 77.5 (23.3) | 10.6 | 23.0 | 49.1 | 77.2 (24.5) | 16.7 | 26.3 | 49.1 |
| 7.1-14.0 | 320 (18.6) | 78 (17.7) | 79.3 (22.9) | 8.4 | 20.0 | 43.8 | 80.0 (21.3) | 3.8 | 19.2 | 42.3 |
| > 14.0 | 194 (11.3) | 41 (9.3) | 79.0 (24.8) | 11.3 | 21.1 | 44.3 | 85.9 (26.9) | 2.4 | 17.1 | 39.0 |
| Current smoker |  |  |  |  |  |  |  |  |  |  |
| No | 1716 (96.7) | 426 (96.4) | 77.6 (23.1) | 9.8 | 22.4 | 48.4 | 77.3 (23.4) | 11.7 | 25.8 | 50.0 |
| Yes | 58 (3.3) | 16 (3.6) | 71.8 (35.0) | 29.3 | 44.8 | 60.3 | 80.9 (32.0) | 18.8 | 31.3 | 43.8 |
| Living alone |  |  |  |  |  |  |  |  |  |  |
| No | 1438 (81.0) | 376 (84.1) | 77.6 (22.9) | 9.7 | 22.0 | 48.7 | 78.0 (23.6) | 11.7 | 25.0 | 48.7 |
| Yes | 338 (19.0) | 71 (15.9) | 76.1 (25.5) | 13.6 | 27.2 | 48.5 | 75.5 (24.5) | 12.7 | 29.6 | 52.1 |
| Self-rated quality of life |  |  |  |  |  |  |  |  |  |  |
| Excellent | 469 (26.4) | 113 (25.5) | 78.0 (22.3) | 7.0 | 21.5 | 49.5 | 79.1 (24.6) | 11.5 | 25.7 | 50.4 |
| Very good | 823 (46.3) | 224 (50.5) | 77.9 (23.0) | 10.2 | 22.1 | 47.0 | 76.1 (23.4) | 12.5 | 25.9 | 49.1 |
| Good, fair or poor | 484 (27.3) | 107 (24.1) | 76.0 (25.5) | 14.0 | 26.0 | 51.0 | 79.2 (23.3) | 10.3 | 25.2 | 47.7 |
| Total intake of vitamin D (IU/day)<sup>3</sup> |  |  |  |  |  |  |  |  |  |  |
| < 100 | 301 (16.9) | 97 (21.7) | 72.9 (22.1) | 15.0 | 27.9 | 57.1 | 73.7 (22.0) | 15.5 | 33.0 | 50.5 |
| 100 to < 200 | 708 (39.8) | 181 (40.6) | 74.8 (23.0) | 13.0 | 28.0 | 53.2 | 74.9 (24.0) | 14.9 | 29.3 | 54.7 |
| 200 to < 500 | 434 (24.4) | 97 (21.7) | 76.6 (23.5) | 8.8 | 22.4 | 49.5 | 77.6 (23.1) | 8.2 | 24.7 | 52.6 |
| ≥ 500 | 334 (18.8) | 71 (15.9) | 87.9 (23.1) | 2.7 | 9.3 | 30.8 | 90.1 (22.9) | 4.2 | 8.5 | 28.2 |

<sup>1</sup> Each observation corresponds to a single blood sample. Training dataset: 1408, 166, and 16 people provided one, two and three blood samples respectively. Validation dataset: 347, 47, and 2 people provided one, two and three blood samples respectively. Missing data in the training dataset: physical activity, 569; time outdoors, 558; tanning ability, 7; body mass index, 59; alcohol consumption, 72; current smoker, 14; living alone, 12; self-rated quality of life, 12; total intake of vitamin D, 11. Missing data in the validation dataset: physical activity, 135; time outdoors, 136; tanning ability, 5; body mass index, 21; alcohol consumption, 6; current smoker, 5; self-rated quality of life, 3; total intake of vitamin D, 1. <sup>2</sup> Month-specific average erythemal daily dose at time of blood draw. It is included as a continuous variables in boosted regression tree models. For simplicity, we have presented summary statistics within categories of ultraviolet irradiation.
<sup>3</sup> Sum of intake from diet and supplements.
Abbreviations: IU, international units; MET, metabolic equivalent of task; SD, standard deviation.

#### Annual surveys

Every 12 months all participants were asked to complete a survey, including those who had withdrawn from the trial. These surveys asked people to rate their QoL using the same five-point scale from the baseline survey. Annual surveys 1, 3 and 5 asked about current weight, allowing us to calculate updated measures of BMI at three points in time. Every annual survey asked questions about the use of supplements containing vitamin D. We used this information to estimate daily intake of vitamin D from off-trial supplements for each year of the intervention.

#### Use of off-trial supplements

To be eligible for the trial, people had to report taking < 500 IU/day vitamin D from supplements. Following randomisation, participants were permitted to increase their daily intake of non-trial supplements up to a maximum of 2,000 IU and still remain in the trial. In addition to capturing supplement use via annual surveys, we also requested that people contact us by phone or online to report any changes to their intake of off-trial supplementary vitamin D.

#### Blood questionnaire

From August 2017, people who gave a blood sample were also asked to complete an additional questionnaire (hereafter called the ‘blood questionnaire’). Using the same wording as at baseline, we asked people to report their skin tanning ability, physical activity, and time spent outdoors during the week prior to having blood taken.

#### Ambient ultraviolet radiation

Ambient ultraviolet (UV) radiation estimates were obtained from the Ozone Monitoring Instrument (OMI) satellite data.^8^ We obtained erythemally weighted average daily UV radiation from the OMUVB product from January 2012 to December 2017. To obtain an estimate of ambient UV radiation at a participant’s place of residence at baseline or time of blood collection, we mapped participants’ residential postcodes to a 1º latitude by 1º longitude grid. We then mapped this to calendar month-specific average erythemal daily doses. For blood samples collected from 2018 onwards, we used monthly averages from 2017.

#### Measurement of serum 25(OH)D concentration

To monitor differences in 25(OH)D distributions between the trial arms, we measured serum 25(OH)D concentrations in a random sample of people. Each year, starting one year after baseline, approximately 350 people from each trial arm received with their annual survey an invitation to provide a blood sample. Those who consented were instructed to attend their local pathology collection centre; a fasting blood sample was collected and sent by courier to our laboratory for initial processing. Samples were collected between March 2015 and February 2020. The 25(OH)D assay was performed using a tandem liquid chromatography mass spectroscopy assay at Metabolomics Australia at the University of Western Australia (a participant in the international Vitamin D Standardization Program).^9,10^

### Data management

#### Training and validation datasets

In the current study we used data and serum 25(OH)D concentrations from the subset of participants who gave a blood sample and were randomised to the placebo arm of the study (maintaining blinding; see below). We partitioned this dataset into training (80%) and validation (20%) datasets. We derived our prediction models in the training dataset and then tested their performance in the validation dataset (details below). A small number of people (11.6%) provided more than one blood sample over the course of the intervention; we ensured that repeated measures from a participant were all allocated to either the training or the validation dataset.

Since blood samples were collected throughout the year we deseasonalised serum 25(OH)D concentrations to account for seasonal variations. We first fitted a sinusoidal model to the measured serum 25(OH)D concentrations (where the sine and cos terms were functions of month of blood collection), and then added the overall mean serum 25(OH)D concentration to the residuals from the sinusoidal model. We dichotomised the deseasonalised serum 25(OH)D concentrations using cut-points of 50 nmol/L, 60 nmol/L and 75 nmol/L, with lower (<) concentrations in each case being the outcome of interest. The Institute of Medicine in the United States concluded that 50 nmol/L is sufficient to ensure bone health in almost all people,^11^ and this is the minimum concentration recommended by The Royal College of Pathologists of Australasia.^12^ We used the cut-point of 75 nmol/L because it is the target concentration recommended by The Endocrine Society;^13^ it is also the approximate median of our measured serum 25(OH)D concentrations. The intermediate cut-point of 60 nmol/L, the approximate first quartile, was included primarily so that we could evaluate how model performance varied with changing cut-points, and because the number <50 nmol/L is potentially too low for effective subgroup analyses. The small number of measured serum 25(OH)D concentrations < 30 nmol/L precluded the use of this as a cut-point.

The dataset included thirteen predictors, four of which were ascertained at baseline only (sex, alcohol consumption, current smoker, and living alone). Tanning ability, physical activity, and time outdoors were taken primarily from the blood questionnaire, which we started asking participants to complete sixteen months after we commenced blood collection. If tanning ability was missing, we used the response at baseline. For people who provided a blood sample at year one (prior to the instigation of the blood questionnaire), we used their levels of physical activity and time outdoors as reported on the baseline survey under the assumption that behaviour would be similar over that time frame. We derived QoL and BMI from the annual survey response prior to the date of blood collection. Since we did not ask about weight on the second or fourth annual surveys, we estimated BMI at these times as follows: if a participant’s BMI in the years prior and subsequent to the year in which they gave blood differed by < 5 kg/m^2^, we used the average of those values; if we didn’t have values for both years, or the difference was ≥ 5 kg/m^2^, we left BMI missing. To calculate total daily intake of vitamin D, we estimated daily intake from supplements using the annual survey that was completed at the time of blood collection, and added this to the reported dietary intake from the baseline survey. We determined the month and participant’s age category at the time of blood collection. Ambient UV radiation was assigned based on the month and place of residence at the time blood was collected. Supplementary Figure 1 summarises the sources of predictor variables and the timing of their ascertainment relative to the time of blood collection.

#### Maintenance of blinding

To remain blinded to treatment allocation, D-Health researchers did not have access to participants’ serum 25(OH)D concentration data. Instead, an external researcher with access to the 25(OH)D data prepared our dataset. They identified which group was receiving the placebo by inspecting summary statistics for serum 25(OH)D concentrations within study arms. The subset of data for the placebo group, including serum 25(OH)D concentration and all potential predictor variables, was then returned to the D-Health team without the randomisation code or the participant identification codes included.

### Data analysis

#### Model fitting

We fitted boosted regression tree (BRT)^14-16^ models to dichotomised deseasonalised serum 25(OH)D concentrations (i.e. used a Bernoulli loss function) for 36 combinations of parameter settings (learning rates of 0.01, 0.005 and 0.001; tree complexities of 2, 3, 4 and 5; and bag fractions of 0.5, 0.6 and 0.7). We used 10-fold cross-validation to determine the optimal number of trees for each choice of parameters. As recommended,^14^ we discarded parameter combinations for which optimal prediction required fewer than 1000 trees. The model with minimum cross-validated deviance was selected as the final model.

#### Assessing the effect of predictors

We estimated the relative contributions of predictors to the final model. For each predictor, we generated a partial dependence plot, showing the marginal effect of the predictor on the response;^15^ these are created by first “averaging” out the effect of all predictors except the one of interest, and then plotting the average fitted value (i.e. the fitted function) against the predictor of interest.

#### Evaluation using the validation dataset

To evaluate each model’s performance, we used it to generate predictions for the validation dataset. We plotted the receiver operating characteristic (ROC) curve, calculated the area under the ROC curve (AUC) and used bootstrapping to estimate a 95% confidence interval (CI) for the AUC. We assessed calibration by plotting observed prevalences against the predicted probabilities for predicted probability ranges defined by deciles. We also calculated the root-mean-squared error (RMSE).

For each model, we plotted sensitivity, specificity, positive predictive value (PPV) and negative predictive value (NPV) versus probability thresholds (used to dichotomise the predictions). We identified the optimal probability threshold using the Youden index, and estimated the PPV, and NPV, and their 95% Agresti-Coull CIs.^17^

#### Predictions for all D-Health participants

We applied the final models to baseline data for all trial participants to predict the probability that each person’s deseasonalised baseline serum 25(OH)D concentration was < 50 nmol/L, < 60 nmol/L, and < 75 nmol/L prior to taking any study tablets. We dichotomised the predictions using the optimal probability threshold identified using the validation dataset.

#### Software

We used SAS version 9.4 (SAS Institute, Inc, Cary, NC) to construct datasets, and to produce summary statistics. R version 3.6.1 was used to fit BRTs, to generate deseasonalised predictions for all D-Health participants, and to produce figures.^18^ We used the gbm package^19^ and additional scripts that were developed for fitting BRTs (gbm.step, gbm.plot, gbm.interactions).^14^ We used the ROCR,^20^ pROC^21^ and modEvA^22^ packages to evaluate models, and the OptimalCutpoints^23^ package to determine the probability threshold to use for dichotomising predictions.

## RESULTS

### Measured serum 25(OH)D concentrations

After excluding one record with an extreme serum 25(OH)D concentration (265 nmol/L), the dataset of serum 25(OH)D concentrations from people randomised to placebo included 2235 observations from 1986 people. Most people (N= 1755 (88.4%)) provided one blood sample only; 213 (10.7%) and 18 (0.9%) people provided two and three blood samples, respectively.

The mean serum 25(OH)D concentration was 77.4 nmol/L (standard deviation (SD) 24.9 nmol/L) (Supplementary Figure 2A). Thirteen percent (n = 291), 24.2% (n = 541) and 48.3% (n = 1079) of samples had serum 25(OH)D concentrations < 50 nmol/L, < 60 nmol/L, and < 75 nmol/L, respectively; only 2.0% (n = 44) had concentrations < 30 nmol/L. Deseasonalising the data reduced slightly the variability in concentrations (SD 23.6 nmol/L) and the percentage with concentrations <50 nmol/L (Supplementary Figure 2B); 10.7%, 23.6%, and 48.9% of deseasonalised concentrations were < 50 nmol/L, < 60 nmol/L, and < 75 nmol/L, respectively.

### Training and validation data

The training and validation datasets had 1788 and 447 samples, respectively. Table 1 shows the distribution of deseasonalised serum 25(OH)D concentration for all participants and by levels of predictor variables. The datasets had very similar percentages of samples with ‘lower’ deseasonalised serum 25(OH)D concentrations (training: 10.5% < 50 nmol/L, 23.1% < 60 nmol, and 48.8 < 75 nmol/L; validation: 11.9% < 50 nmol/L, 25.7% < 60 nmol/L, and 49.2% < 75 nmol/L).

### Final parameter settings

The models for < 50 nmol/L, < 60 nmol/L, and < 75 nmol/L used tree complexities of 5, 2, and 5, respectively, learning rates of 0.001, 0.001, and 0.005, respectively, and bag fractions of 0.5, 0.5, and 0.6, respectively, and their cross-validated deviance was minimised using 3850, 3850, and 1000 trees (Supplementary Tables 1 to 3).

### Associations with predictors

The two most influential predictors in all models were ambient UV radiation and total intake of vitamin D from food and supplements (Table 2). The interaction between these two variables was always amongst the top two strongest two-way interactions (Supplementary Table 4). Partial dependency plots for these predictors are shown in Figure 1. As expected, the predicted probability of ‘lower’ serum 25(OH)D concentration decreased with increasing total intake of vitamin D. There was a non-linear relationship with increasing UV radiation, decreasing steeply before becoming relatively stable. Partial dependency plots for the predictors that were ranked 3^rd^ to 13^th^ (least) influential are shown in Supplementary Figures 3 to 5.

**Figure 1.**
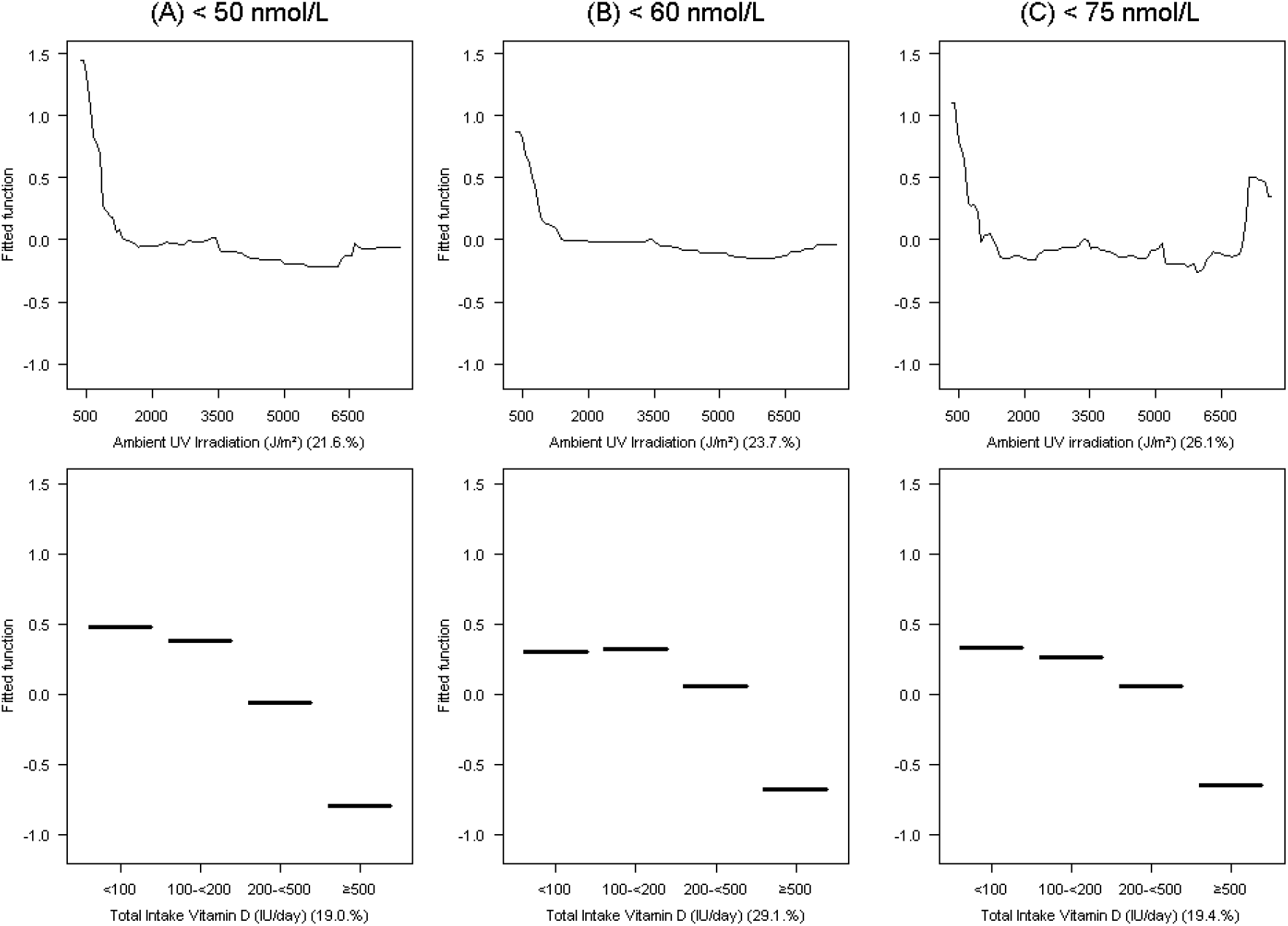
Partial dependence plots^1^ for the two most influential^2^ variables (ambient UV irradiation^3^ and total intake of vitamin D^4^) in the boosted regression tree models^5^ for deseasonalised serum 25(OH)D concentration (A) < 50 nmol/L, (B) < 60 nmol/L, and (C) < 75 nmol/L. ^1^ Fitted functions have been centered by subtracting their mean. ^2^ The relative contribution of each predictor variable is given in brackets. ^3^ Month-specific average erythemal daily dose at time of blood draw. ^4^ Sum of intake from diet and supplements. ^5^ The models were fitted to a training dataset containing n = 1788 blood samples. The models for < 50 nmol/L, < 60 nmol/L, and < 75 nmol/L used tree complexities of 5, 2, and 5, respectively, learning rates of 0.001, 0.001, and 0.005, respectively, and bag fractions of 0.5, 0.5, and 0.7, respectively, and their cross-validated deviance was minimised using 3850, 3850, and 1000 trees, respectively.

**Table 2.** Relative contributions (%) of predictor variables in the boosted regression tree models for deseasonalised serum 25(OH)D concentration < 50 nmol/L, < 60 nmol/L, and < 75 nmol/L.

| < 50 nmol/L |  | < 60 nmol/L |  | < 75 nmol/L |  |
| --- | --- | --- | --- | --- | --- |
| Predictor variable | RC (%) | Predictor variable | RC (%) | Predictor variable | RC (%) |
| Ambient UV radiation<sup>1</sup> | 21.6 | Total intake of vitamin D<sup>2</sup> | 29.1 | Ambient UV radiation<sup>1</sup> | 26.1 |
| Total intake of vitamin D<sup>2</sup> | 19.0 | Ambient UV radiation<sup>1</sup> | 23.7 | Total intake of vitamin D<sup>2</sup> | 19.4 |
| Time outdoors | 11.3 | Time outdoors | 10.3 | Alcohol consumption | 12.9 |
| Alcohol consumption | 8.9 | Current smoker | 6.2 | Time outdoors | 9.6 |
| BMI | 8.4 | QoL | 5.3 | BMI | 7.4 |
| QoL | 6.9 | Alcohol consumption | 5.2 | Month of blood draw | 5.1 |
| Physical activity | 6.0 | Physical activity | 4.2 | Physical activity | 4.5 |
| Month of blood draw | 4.7 | Living alone | 4.2 | Tanning ability | 4.3 |
| Current smoker | 4.5 | BMI | 4.2 | Age | 4.0 |
| Tanning ability | 3.8 | Tanning ability | 3.8 | QoL | 2.6 |
| Living alone | 1.9 | Month of blood draw | 1.8 | Sex | 1.6 |
| Age | 1.7 | Age | 1.8 | Current smoker | 1.5 |
| Sex | 1.2 | Sex | 0.4 | Living alone | 1.1 |

<sup>1</sup> Month-specific average erythral daily dose at time of blood draw. <sup>2</sup> From diet and supplements Abbreviations: BMI, body mass index; QoL, quality of life; RC, relative contribution; UV, ultraviolet

### Evaluation using validation data

The models for < 50 nmol/L and < 60 nmol/L had very similar discriminatory ability, with AUCs of 0.71 (95% CI 0.63–0.78) and 0.70 (95% CI 0.65–0.76), respectively (Figure 2). The AUC for the model predicting < 75 nmol/L was slightly lower (0.66, 95% CI 0.61–0.71). All models were reasonably well calibrated (Figure 3). The RMSE was 0.31, 0.42, and 0.48 for the models for < 50 nmol/L, < 60 nmol, and < 75 nmol/L, respectively.

**Figure 2.**
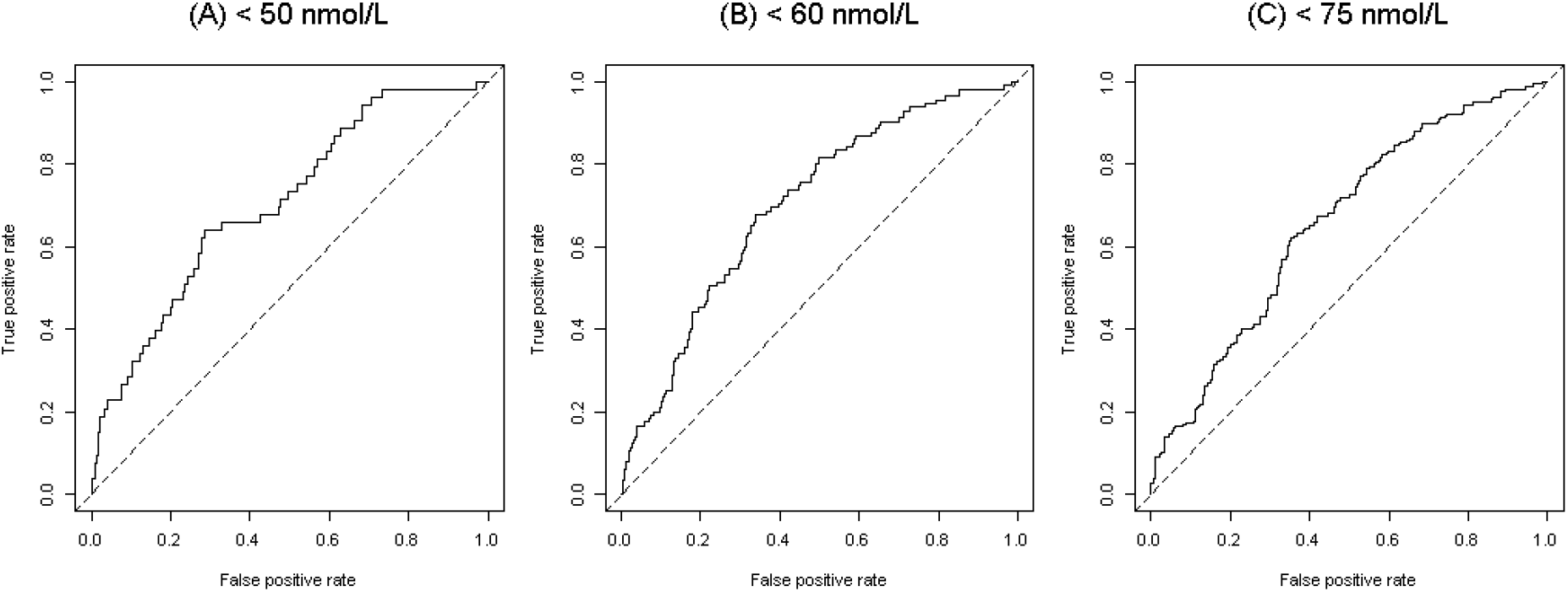
Receiver operating characteristic curves for the boosted regression tree models for deseasonalised serum 25(OH)D concentration (A) < 50 nmol/L, (B) < 60 nmol/L, and (C) < 75 nmol/L; models applied to the validation dataset (n = 447).

**Figure 3.**
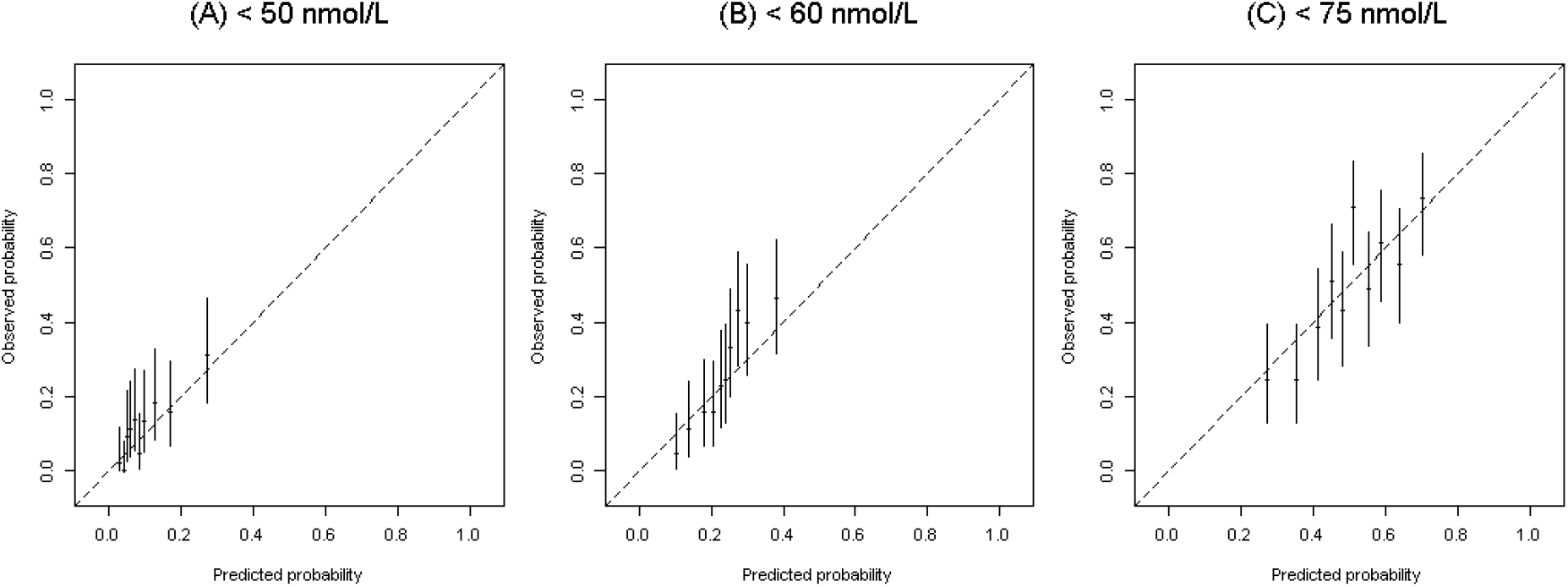
Calibration plots^1^ for the boosted regression tree models for deseasonalised serum 25(OH)D concentration (A) < 50 nmol/L, (B) < 60 nmol/L, and (C) < 75 nmol/L; models applied to the validation dataset (n = 447). ^1^ Predicted probability ranges defined by deciles.

Supplementary Figure 6 shows the sensitivity, specificity, PPV, and NPV plotted against probability threshold for the model for < 50 nmol/L. The model had very high NPV across a wide range of thresholds but its PPV was generally poor. Using the optimal threshold to dichotomise predictions, the NPV was excellent (0.94, 95% CI 0.90–0.96), the sensitivity (0.64, 95% CI 0.51–0.76) and specificity (0.72, 95% CI 0.70–0.76) were moderately good, and the PPV was poor (0.23, 95% CI 0.17–0.31) (Table 3). Compared with the model for < 50 nmol/L, the model for < 60 nmol/L had slightly better PPV, its NPV was very good, and its sensitivity and specificity were similar (Table 3, Supplementary Figure 7). All performance measures for the model predicting < 75 nmol/L were between 0.62 and 0.65, inclusive (Table 3, Supplementary Figure 8).

**Table 3.** Sensitivity, specificity, positive predictive value (PPV), and negative predictive value (NPV) of boosted regression tree models^1^ when predictions are dichotomised using the optimal thresholds;^2^ values calculated using the validation dataset (n = 447).

| Model<sup>1</sup> | Probability threshold | Estimate (95% CI)<sup>3</sup> |  |  |  |
| --- | --- | --- | --- | --- | --- |
|  |  | Sensitivity | Specificity | PPV | NPV |
| < 50 nmol/L | 0.103 | 0.642 (0.507, 0.757) | 0.716 (0.669, 0.758) | 0.233 (0.172, 0.308) | 0.937 (0.904, 0.959) |
| < 60 nmol/L | 0.244 | 0.678 (0.588, 0.757) | 0.663 (0.610, 0.711) | 0.411 (0.343, 0.482) | 0.856 (0.808, 0.894) |
| < 75 nmol/L | 0.501 | 0.623 (0.557, 0.684) | 0.648 (0.583, 0.707) | 0.631 (0.565, 0.693) | 0.639 (0.575, 0.698) |

<sup>1</sup> Modelling deseasonalised serum 25(OH)D concentration < 50 nmol/L, < 60 nmol/L, and < 75 nmol/L. <sup>2</sup> Value identified using the Youden index.<sup>3</sup> CIs estimated using the method proposed by Agresti and Coull.
Abbreviation: CI, confidence interval.

### Predictions for all participants at baseline

We applied the models to baseline data for all D-Health Trial participants and dichotomised predictions using the probability thresholds in Table 3. Using the model for < 50 nmol/L, 24.4% of people were predicted to have deseasonalised serum 25(OH)D concentration < 50 nmol/L prior to taking any study tablets. The percentage < 60 nmol/L was 30.1%, and just over one third (35.6%) of people were predicted to have a concentration < 75 nmol/L. Supplementary Table 5 shows a cross-classification of predictions from the three models.

The percentage of people with ‘lower’ predicted serum 25(OH)D concentration varied greatly with total intake of vitamin D, ranging from <2% amongst people whose total intake was ≥ 500 IU/day to >40% in those taking < 100 IU/day (Table 4). Those with greater physical activity or more time outdoors were less likely to have low predicted 25(OH)D concentration. People with BMI ≥ 30 kg/m^2^, or who lived alone, had poorer QoL, or smoked were more likely to be predicted to have lower deseasonalised serum 25(OH)D concentration than people who were not in these groups.

**Table 4.** Percentage of participants with predicted deseasonalised serum 25(OH)D concentration < 50 nmol/L, < 60 nmol/L, and < 75 nmol/L prior to taking any study tablets by levels of predictor variables; predictions obtained by applying boosted regression tree models^1^ to baseline data for all participants of the D-Health Trial.

| Predictor variable | N (%)<sup>2</sup> | Predicted deseasonalised serum 25(OH)D concentration |  |  |
| --- | --- | --- | --- | --- |
|  |  | % < 50 nmol/L | % < 60 nmol/L | % < 75 nmol/L |
| **Age (years)** |  |  |  |  |
| < 70 | 11086 (52.0) | 25.1 | 28.7 | 27.5 |
| 70 to < 75 | 5796 (27.2) | 23.0 | 29.8 | 40.8 |
| $\geq 75$ | 4428 (20.8) | 24.5 | 33.8 | 49.1 |
| **Sex** |  |  |  |  |
| Men | 11530 (54.1) | 21.8 | 27.1 | 37.8 |
| Women | 9780 (45.9) | 27.5 | 33.5 | 33.0 |
| **Month of randomisation** |  |  |  |  |
| January | 814 (3.8) | 20.4 | 22.1 | 37.6 |
| February | 1582 (7.4) | 19.3 | 20.0 | 27.3 |
| March | 1517 (7.1) | 23.9 | 29.1 | 33.1 |
| April | 1433 (6.7) | 25.6 | 33.3 | 34.0 |
| May | 1419 (6.7) | 36.1 | 46.5 | 41.6 |
| June | 1298 (6.1) | 50.6 | 61.9 | 61.6 |
| July | 1596 (7.5) | 46.4 | 58.0 | 58.1 |
| August | 1675 (7.9) | 25.6 | 35.8 | 32.9 |
| September | 1948 (9.1) | 22.9 | 30.5 | 31.7 |
| October | 2557 (12.0) | 17.5 | 23.0 | 33.0 |
| November | 2839 (13.3) | 13.2 | 14.9 | 26.8 |
| December | 2632 (12.4) | 14.9 | 15.2 | 29.3 |
| **Ambient ultraviolet radiation (J/m<sup>2</sup>)<sup>3</sup>** |  |  |  |  |
| < 1250 | 3338 (15.7) | 50.5 | 62.9 | 64.9 |
| 1250 to < 2500 | 4102 (19.2) | 23.6 | 31.8 | 26.5 |
| 2500 to < 3750 | 3268 (15.3) | 23.5 | 31.6 | 36.6 |
| 3750 to < 5000 | 3861 (18.1) | 17.4 | 21.4 | 30.0 |
| $\geq 5000$ | 6741 (31.6) | 16.4 | 17.0 | 29.4 |
| **Physical activity (METs/week)** |  |  |  |  |
| Low (< 18) | 6889 (33.4) | 44.5 | 45.5 | 46.3 |
| Moderate (18 to < 45) | 6835 (33.1) | 14.7 | 27.2 | 33.6 |
| High ($\geq 45$) | 6902 (33.5) | 11.7 | 15.3 | 24.9 |
|  |  | % < 50 nmol/l | % < 60 nmol/l | % < 75 nmol/l |
| Time outdoors (hours/week) |  |  |  |  |
| < 5 | 4066 (19.6) | 58.6 | 51.0 | 46.5 |
| 5 to < 10 | 4240 (20.4) | 25.2 | 39.8 | 44.7 |
| 10 to < 15 | 3763 (18.1) | 15.8 | 34.9 | 42.1 |
| 15 to < 25 | 4548 (21.9) | 11.9 | 13.5 | 23.6 |
| ≥ 25 | 4157 (20.0) | 8.9 | 10.2 | 19.7 |
| Tan moderately or deeply |  |  |  |  |
| No | 6929 (32.9) | 35.6 | 42.5 | 50.6 |
| Yes | 14155 (67.1) | 18.8 | 23.9 | 28.3 |
| Body mass index (kg/m²) |  |  |  |  |
| < 25 | 6417 (30.3) | 22.5 | 23.7 | 20.3 |
| 25 to < 30 | 9029 (42.6) | 19.6 | 25.6 | 37.8 |
| ≥ 30 | 5745 (27.1) | 33.6 | 44.2 | 49.7 |
| Alcohol consumption (drinks/week) |  |  |  |  |
| < 1.0 | 5046 (24.6) | 29.9 | 40.0 | 52.7 |
| 1.0-2.0 | 3252 (15.9) | 24.0 | 32.3 | 37.0 |
| 2.1-7.0 | 5852 (28.6) | 23.1 | 27.4 | 37.2 |
| 7.1-14.0 | 3754 (18.3) | 18.0 | 20.2 | 19.5 |
| > 14.0 | 2580 (12.6) | 21.6 | 22.7 | 20.2 |
| Current smoker |  |  |  |  |
| No | 20229 (95.8) | 21.9 | 27.9 | 34.8 |
| Yes | 896 (4.2) | 79.9 | 78.0 | 55.8 |
| Living alone |  |  |  |  |
| No | 16960 (80.0) | 22.4 | 26.6 | 34.8 |
| Yes | 4233 (20.0) | 31.8 | 43.5 | 38.6 |
| Self-rated quality of life |  |  |  |  |
| Excellent | 4359 (20.9) | 13.6 | 20.0 | 30.4 |
| Very good | 9601 (46.1) | 22.2 | 25.9 | 32.6 |
| Good, fair or poor | 6855 (32.9) | 33.9 | 42.1 | 42.8 |
| Total intake of vitamin D (IU/day)<sup>4</sup> |  |  |  |  |
| < 100 | 3866 (18.2) | 41.5 | 45.8 | 56.4 |
| 100 to < 200 | 8872 (41.8) | 33.0 | 43.0 | 46.0 |
| 200 to < 500 | 5151 (24.3) | 12.2 | 15.3 | 24.3 |
| ≥ 500 | 3321 (15.7) | 0.2 | 0.0 | 1.3 |

<sup>1</sup> The models were fitted to a training dataset containing n = 1788 blood samples. The models for < 50 nmol/L, < 60 nmol/L, and < 75 nmol/L used tree complexities of 5, 2, and 5, respectively, learning rates of 0.001, 0.001, and 0.005, respectively, and bag fractions of 0.5, 0.5, and 0.7, respectively, and their cross-validated deviance was minimised using 3850, 3850, and 1000 trees, respectively. <sup>2</sup> Missing data: physical activity, 684; time outdoors, 536; tanning ability, 226; body mass index, 119; alcohol consumption, 826; current smoker, 185; living alone, 117; self-rated quality of life, 495; total intake of vitamin D, 100. <sup>3</sup> Month-specific average erythemal daily dose at time of randomisation. Ambient ultraviolet radiation was included as a continuous variables in boosted regression tree models. For simplicity, we have presented summary statistics within categories of ultraviolet radiation. <sup>4</sup> Total intake from diet and supplements.

## DISCUSSION

To enable future exploratory analyses stratified by baseline vitamin D status, we developed BRT models to predict whether or not each participant in the D-Health Trial had ‘low’ deseasonalised serum 25(OH)D concentration prior to taking any study tablets. Separate models were fitted to predict concentrations < 50 nmol/L, < 60 nmol/L, and < 75 nmol/L, and all identified ambient UV radiation and total intake of vitamin D from food and supplements as the strongest predictors of a ‘low’ serum 25(OH)D concentration. The discriminatory ability of the models for < 50 nmol/L and < 60 nmol/L were very similar; it was slightly lower for the model for < 75 nmol/L. Approximately one quarter of participants had predicted deseasonalised serum 25(OH)D concentration < 50 nmol/L (24.4%), and the percentages of participants who had predicted concentrations < 60 nmol/L and < 75 nmol/L were 30.1% and 35.6%, respectively.

In the 2011-2013 Australian Health Survey (AHS) 15% of people aged 65 to 74 years, and 20% of people aged ≥ 75 years had serum 25(OH)D concentration < 50 nmol/L.^24^ These values are somewhat higher than the 13% (non-deseasonalised) we observed in blood samples from people randomised to placebo. Similarly, 48% of our samples had serum 25(OH)D concentration < 75 nmol/L compared with the 60% reported for older people (aged ≥ 55 years) in a recent analysis of determinants of vitamin D deficiency in the general Australian population using the AHS data.^25^ The AHS used a nationally representative sample and its estimates are survey-weighted to the Australian population (excluding people who live in very remote areas).^26^ The percentages we present are unweighted, so discordant estimates might reflect differences in residential location or ethnicity (91% of our participants described their ancestry as British/European).^4^ Moreover, people participating in the D-Health Trial are, on average, healthier than the general older Australian population.^4^

The primary source of vitamin D is sun exposure. Hence, it is not surprising that ambient UV radiation should be an important predictor in our models. The strong contribution of total intake of vitamin D to the model is also consistent with expectations. Obesity and low levels of physical activity have been shown to be independent predictors of vitamin D deficiency in the general Australian population,^25^ and predictions from our model are consistent with these findings. Similarly, the association we observed between smoking and lower predicted serum 25(OH)D concentration was also noted in the AHS.^24^ Genetics account for considerable variability in serum 25(OH)D concentrations,^27^ and this is likely to explain some of the variation not accounted for by our models.

The percentage of participants with predicted deseasonalised concentration < 50 nmol/L was considerably higher than the observed percentage for all blood samples (24.4% versus 10.7%) and this discrepancy might be explained, in part, by differences in supplemental intake of vitamin D at baseline and at the times when we collected blood samples. To be eligible to participate, people needed to be taking ≤ 500 IU/day supplementary vitamin D; after randomisation (when blood samples were collected) people could take ≤ 2000 IU/day off-trial supplementary vitamin D without having to withdraw. However, we discouraged the use of high-dose supplementation, and the percentage consuming ≥ 500 IU/day from food and supplements was not greatly higher in the training dataset (i.e. at time of blood draw) to at baseline (18.8% versus 15.7%).

The model predicting 25(OH)D concentration < 50 nmol/L had an excellent NPV and poor PPV, both of which are partly explained by the low prevalence of samples with concentration < 50 nmol/L. The low PPV will account for some of the discrepancy between the observed and predicted percentages. This has implications for analyses we perform stratified by this variable since there will be a considerable number of people classified incorrectly as having deseasonalised serum 25(OH)D concentration < 50 nmol/L which will reduce the probability that we will detect an actual effect (benefit or otherwise) of vitamin D supplementation amongst people with lower serum 25(OH)D. Conversely, if an interaction between randomisation and baseline serum 25(OH)D concentration is significant in future analyses, we can be reasonably confident that the actual effect is greater than estimated.

The poor PPV of the model for < 50 nmol/L highlights that predicting low concentrations is intrinsically difficult. Since the model for < 60 nmol/L had similar discriminatory power as the model for < 50 nmol/L, we may perform exploratory analyses of D-Health outcomes stratified using a cut-point of 60 nmol/L. In contrast, the predictions from the model for < 75 nmol/L are not sufficiently precise to use for future analyses.

There are trade-offs inherent in predicting rather than measuring baseline vitamin D status. Any future analyses that are stratified by predicted deseasonalised serum 25(OH)D concentration must be considered exploratory. Some misclassification of true baseline vitamin D status is inevitable. We have relied heavily on self-reported data. Although we cleaned data during the course of the intervention, often phoning participants to check questionable survey responses, errors are likely to remain. Some data were not collected at the time blood was provided and we assumed that the rankings of time outdoors and physical activity were relatively stable during the year after randomisation. Likewise, we assumed stability of dietary habits. The UV radiation data are not ground-level measurements and, for blood samples collected from 2018 onwards, we used data from 2017. However, the relative ranking of areas is unlikely to have changed substantively, particularly since we used monthly average values. Further, since the machine learning algorithm implicitly handles any non-linear but one-to-one transformation from nominal to true values, the use of older data should not have had a deleterious effect on our modelling.

The merits of using BRT models have been enumerated elsewhere.^14^ We used this approach because: we could use all serum 25(OH)D concentrations regardless of whether some of the predictor variables had missing values with no need for imputation; we did not have to specify interactions; and we could introduce stochasticity through the use of a bag fraction. The latter is likely to improve the performance of the prediction model when applied to the entire cohort. Unlike a traditional single tree model, the BRT model is also less prone to over-fitting the training data.

Motivated by the prohibitive cost of collecting blood samples from all participants in a large field trial of more than 20,000 participants, we have exploited compliance monitoring data to develop models to predict serum 25(OH)D concentration. These models are intended to generate predictions for the D-Health Trial so that we may perform exploratory analyses stratified by the predicted baseline vitamin D status. However, the general approach we have outlined may also prove useful in other trial settings where there is an obstacle to exhaustive data collection.

## Data Availability

The data are not publicly available.

## Abbreviations

25(OH)D: 25 hydroxy vitamin D
AHS: Australian Health Survey
AUC: area under the ROC curve
BMI: body mass index
BRT: boosted regression tree
CI: confidence interval
IU: international units
MET: metabolic equivalent of task
NPV: negative predictive value
PPV: positive predictive value
QoL: quality of life
RCT: randomised controlled trial
RMSE: root-mean-squared error
ROC: receiver operating characteristic
SD: standard deviation
UV: ultraviolet.

## Clinical Trial Registration Number

Australian New Zealand Clinical Trials Registry: ACTRN12613000743763. Registered on 4 July 2013.

## Acknowledgements

We would like to acknowledge the D-Health Trial staff and members of the Data and Safety Monitoring Board (Patricia Valery, Ie-Wen Sim, Kerrie Sanders). We also extend our thanks to the D-Health Trial participants who committed to this research.

## Grants and fellowships

The D-Health Trial is funded by National Health and Medical Research Council (NHMRC) project grants (APP1046681, APP1120682). PM Webb and DC Whiteman are supported by fellowships from the NHMRC (GNT1173346, APP1155413). DSA McLeod is supported by a Metro North Clinician Research Fellowship and a Queensland Advancing Clinical Research Fellowship. H Pham is supported by a University of Queensland PhD Scholarship. MW Clarke is affiliated to Metabolomics Australia, University of Western Australia, Perth, Western Australia, Australia. Measurement of serum 25(OH)D concentration was supported by infrastructure funding from the Western Australian State Government in partnership with the Australian Federal Government, through Bioplatforms Australia and the National Collaborative Research Infrastructure Strategy (NCRIS).

## CRediT author statement

**Mary Waterhouse:** Conceptualisation, Methodology, Formal analysis, Data Curation, Writing - Original Draft, Writing - Review & Editing, Visualisation. **Catherine Baxter:** Investigation, Data Curation, Project administration. **Briony Duarte Romero:** Investigation, Data Curation, Project administration. **Donald S. A. McLeod:** Writing - Review & Editing. **Dallas R. English:** Conceptualisation, Funding acquisition. **Bruce K. Armstrong:** Conceptualisation, Writing - Review & Editing, Funding acquisition. **Michael W. Clarke:** Resources, Writing - Review & Editing. **Peter R. Ebeling:** Conceptualisation, Writing - Review & Editing, Funding acquisition. **Gunter Hartel:** Writing - Review & Editing. **Michael G. Kimlin:** Conceptualisation, Writing - Review & Editing, Funding acquisition. **Rachel L. O’Connell:** Conceptualisation, Writing - Review & Editing, Funding acquisition. **Hai Pham:** Data Curation. **Rachael M. Rodney Harris:** Resources, Writing - Review & Editing. **Jolieke C. van der Pols:** Conceptualisation, Writing - Review & Editing, Funding acquisition. **Alison J. Venn:** Conceptualisation, Writing - Review & Editing, Funding acquisition. **Penelope M. Webb:** Conceptualisation, Writing - Review & Editing. **David C. Whiteman:** Conceptualisation, Writing - Review & Editing. **Rachel E. Neale:** Conceptualisation, Methodology, Investigation, Data Curation, Writing - Review & Editing, Supervision, Project administration, Funding acquisition.

